# Quantitative form and fit of N95 filtering facepiece respirators are retained and coronavirus surrogate is inactivated after heat treatments

**DOI:** 10.1101/2020.04.15.20065755

**Authors:** Travis Massey, Monica Borucki, Samuel Paik, Kyle Fuhrer, Mihail Bora, Staci Kane, Razi Haque, Salmaan Baxamusa

## Abstract

Re-use of filtering facepiece respirators (FFRs, commonly referred to as N95s) normally meant for single use only is becoming common in healthcare facilities due to shortages caused by the COVID19 pandemic. Here we report that mouse hepatitis virus (MHV) initially seeded on FFR filter material is inactivated (6 log reduction as measured by 50% tissue culture infective dose (TCID50)) after dry heating at 75 ^º^C for 30 minutes. We also find that the quantitative fit of FFRs after heat treatment at this temperature, under dry conditions or at 90% relative humidity, is not affected by single or ten heating cycles. Previous studies have reported that the filtration efficiency of FFR filters is not negatively impacted by these heating conditions. These results suggest that thermal inactivation of coronaviruses is a potentially rapid and widely deployable method to re-use N95 FFRs in emergency situations where re-using FFRs is a necessity and broad-spectrum sterilization is unavailable. However, we also found that a heat source that emits radiation (e.g., an exposed heating element) results in rapid qualitative degradation of the FFR. Finally, we discuss differences in the results reported here and other recent studies investing heat as a means to recycle FFRs and suggest that overall wear time and donning/doffing cycles are important factors that need to be considered.

## Introduction

The worldwide global demand for N95 filtering face respirators (FFRs) for healthcare professionals will quickly outpace supply as the global COVID19 pandemic continues. Several protocols for disinfecting and reusing FFRs that are normally for one-time use only have been proposed,^1^ and the FDA has authorized emergency use of vapor hydrogen peroxide (VHP) as a broad-spectrum sterilant for re-use of FFRs^2^ Because VHP requires centralized operations and specialized equipment, it may not be available in all emergency situations.

In this study, we investigate the use of heat treatment at 75 ºC as a potential method for recycling N95 FFRs. Although not a broad-spectrum sterilant, heat treatments may be easily adapted in a variety of settings and filter efficiency of FFRs has been reported to be maintained after one hour at 90 ºC under dry heat for 1 hour.^3^ A very recent report found that filter efficiency of meltblown polypropylene filter fabric was not affected by either dry or moist heat at ≤85 ºC for up to 50 cycles.^4^ Therefore, the heating schedule used is cool enough to maintain the filter efficiency of the FFR.

In addition to efficiently capturing aerosolized particles in the filter elements, the FFR must also form a good seal around the nose and face of the wearer. Reports are now emerging on the quality of the seal via ***quantitative*** fit testing of FFRs following decontamination protocols. A recent evaluation of typical hospital decontamination protocols showed that quantitative fit was retained following multiple cycles of ethylene oxide, VHP, and (for some FFR models) autoclaving.^5^ Another recent study, released during the preparation of the present report, has reported that quantitative fit factor of FFRs is retained for two cycles of dry heat at 70 ºC but fail thereafter.^6^ In this study, we performed standard OSHA quantitative fit testing on FFRs that underwent both one and ten heating cycles at 75 ºC under both dry and humid conditions.

Although heating at this temperature is not a broad-spectrum sterilant, previous studies in liquid media have reported SARS-CoV-1 inactivation at 60 ºC/30 minutes (≥5 log reduction in viral activity) ^7^ and at 75 ºC/15 minutes (≥4 log reduction).^8^ Most recently, SARS-CoV-2 has been shown to be inactivated (≥6.8 log reduction) by heating to 70 ºC for 5 minutes in liquid media.^9^ While promising, confirmation that the virus can be inactivated after deposition on FFR filter material is needed to investigate FFR re-use. A study released during the preparation of the present report found that SARS-CoV-2 on FFR filter material could be inactivated (≥4 log reduction) after dry heating at 70ºC for 60 minutes.^6^ To simulate decontamination under use conditions, we also measured the activity of a coronavirus (in this case, mouse hepatitis virus (MHV) as a surrogate to SARS-CoV-2), initially inoculated onto FFR filter material and titered using a TCID50 protocol before and after dry heat treatment at 75 ºC.

## Method

### Heat treatments

Prior to heat treatment, a volunteer participant briefly fitted a new, unused FFR (3M Model 8210) to their face and nose structure to simulate a first-time use. FFRs were then loaded into a sterilization pouch (CrossTex Sure-Check, SCL12182).

For dry heating, the FFRs were loaded into an oven (Cascade Tek TFO-1) pre-warmed to 75 ºC and operating at ambient humidity. We estimate that ambient humidity at 75 ºC is <5% relative humidity. The oven door was held open for <30 seconds during loading and experienced a temperature loss of <2 ºC. Some FFRs were instrumented with a thermocouple as shown in Figure 1a to monitor the thermal history. Figure 1b shows the thermal history of several FFRs from both the top and bottom shelf of the oven. The FFRs heated to 75 ºC in approximately 5 minutes and this temperature was maintained to ±3 ºC throughout the 30-minute treatment. FFRs that were subjected to multiple cycles were allowed to cool to room temperature but were not removed from the pouch or re-donned prior to the next heating cycle.

**Figure 1:**
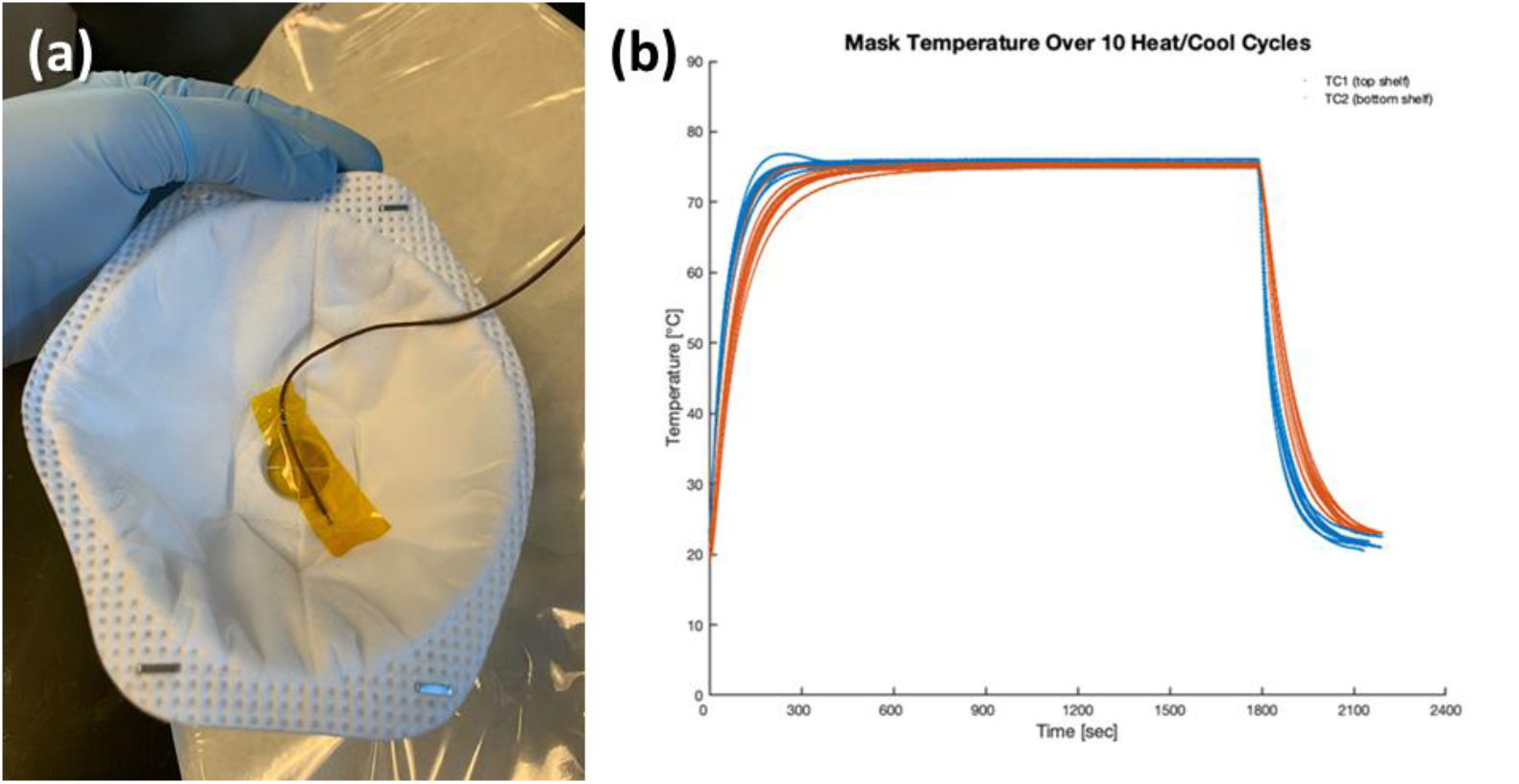
a) Thermocouple attached to an FFR with Kapton tape. This picture shows a 3M Model 8511 N95 FFR used for validating the process. All fit tests were performed with a 3M Model 8210 N95 FFR. b) Thermal history of FFRs on top (blue) and bottom (red) shelf of oven over multiple cycles.

For humid heating, FFRs were loaded into an environmental chamber (Espec SH-242). We visually confirmed that the pouch allowed steam to permeate to the surface of the FFR during experimental process development. The chamber was programmed to ramp over 15 minutes to 75 ºC and 90% relative humidity, hold those conditions for 30 minutes, and ramp back down to room temperature and humidity over 15 minutes. For FFRs treated for multiple cycles, these conditions were held for seven hours before the next heat and humidity cycle began. The FFRs were not re-donned between cycles.

Neither oven used has exposed heating elements, as the infrared radiation emitted from exposed heating elements may be absorbed by the polymer components of the FFR and cause rapid heating and damage to the FFR (see Results and Dscussion).

### Quantitative fit test

Quantitative fit tests were performed per OSHA fit test protocol 1910.134, Appendix A using a TSI PortaCount Respirator Fit Tester 8038 (TSI Instruments, Shoreview MN).^10^ This instrument does not test the effectiveness of the filter, which previous studies have validated up to 90 ºC.^3,4^ The tests performed in this study quantify changes to the sealing surface insofar as their effect on fit. The passing criteria for the fit test was a quantitative fit factor of 100, which is the OSHA criteria for new, unused FFRs.

A sodium chloride (NaCl) aerosol generator and two humidifiers with tunable droplet size set at the smallest droplet size setting were used to achieve background levels of aerosol. The PortaCount Fit Tester 8038 uses a pre-selector to ensure that the detected aerosols reflect those penetrating the sealing surfaces and not the filter itself. Aerosolized particle counts were compared inside and outside the FFR while the participant performed a series of seven 60-second and one 15-second breathing, movement, and speaking exercises including:

- Normal breathing
- Deep breathing
- Head side-to-side
- Head up and down
- Talking
- Grimacing (15 seconds)
- Bending over/reaching down
- Normal breathing

Samples were only fit-tested on the volunteer who originally donned and doffed the FFR prior to the heat treatment. Fit tests on samples 01-08 (Volunteer A) were performed sequentially, but the order of fit tests on samples 09-16 (Volunteer B) was randomized.

A total of 18 FFRs were treated in this study as shown in Table 1. For each heating schedule tested, the samples were cycled either once or ten times. Because the fit test requires inserting a metal rivet into the FFR, we considered the test destructive. Therefore, pre- and post-treatment measurements were not possible on the same FFR and control measurements were instead made for each volunteer with new, unused FFRs without heat treatment.

**Table 1:** Sample set of FFRs and conditions tested in this study.

| Sample | Humidity | Heating cycles | Volunteer |
| --- | --- | --- | --- |
| 01, 02, 03, 04 | Dry | 1x | A |
| 05, 06, 07, 08 | Dry | 10x | A |
| 17 (control) | N/A | None | A |
| 09, 10, 11, 12 | 90% RH | 1x | B |
| 13, 14, 15, 16 | 90% RH | 10x | B |
| 18 (control) | N/A | None | B |

### Viral activity measurements

Twenty microliters of viral stock diluted in Dulbecco’s Modified Eagle Medium (DMEM) media (Thermo Scientific, Waltham MA) with 10% Fetal Bovine Serum (FBS) and antibiotics was inoculated onto 12 replicate ½ cm^2^ coupons cut from an FFR (3M Model 8210). The inocula were dried inside sterile petri plates within a biosafety cabinet for 1 hour, and 2 sets of 3 coupons were placed in 2 autoclave bags (CrossTex Sure-Check, SCL12182), sealed, and placed in the oven pre-heated to 75 ºC (Cascade Tek TFO-1). At 30 minutes one bag containing 3 coupons were was removed from the oven and allowed to cool for 30 minutes, and the second bag containing 3 coupons was removed from the oven at 60 minutes and allowed to cool for 30 minutes. Control coupons were stored at room temperature and were processed at the same timepoints as when the corresponding 30- and 60-minute coupons were processed. Thirty minutes after removal from the oven, each coupon was immersed in 2 ml of media (DMEM, 10% FBS, and antibiotics) and vortexed intermittently for 10 minutes to dislodge the viral particles from the coupon into the media. The virus was then titered by TCID50 assays using 17CL-1 cells. Cytopathic effect for each well was recorded day 3 post-inoculation and TCID50 titer was calculated using the Spearman and Karber method.^11^

## Results and discussion

### Quantitative fit test results

Results of the quantitative fit tests are shown in Table 2. A passing score is 100, and the maximum possible score is 200.

**Table 2:**
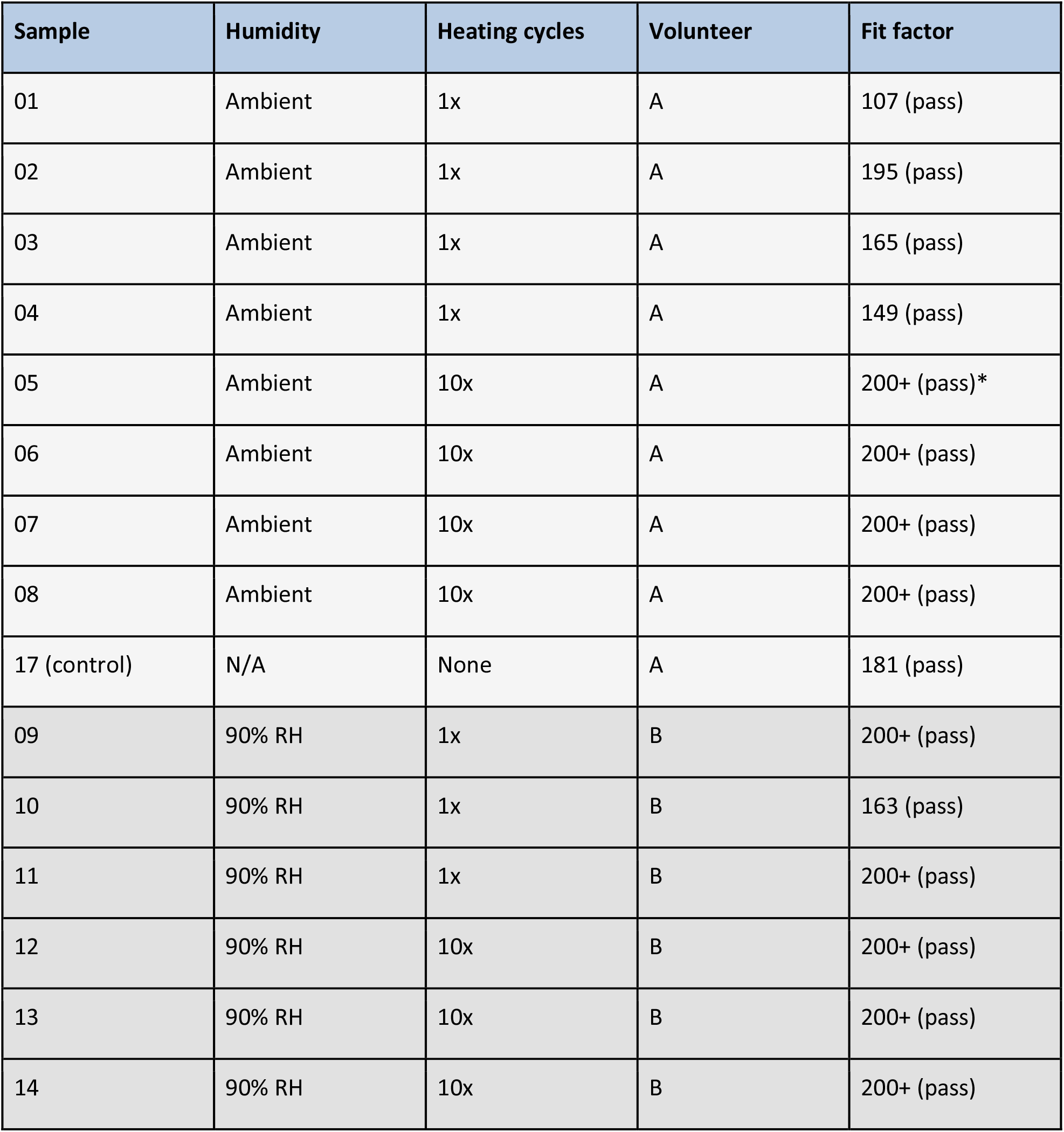
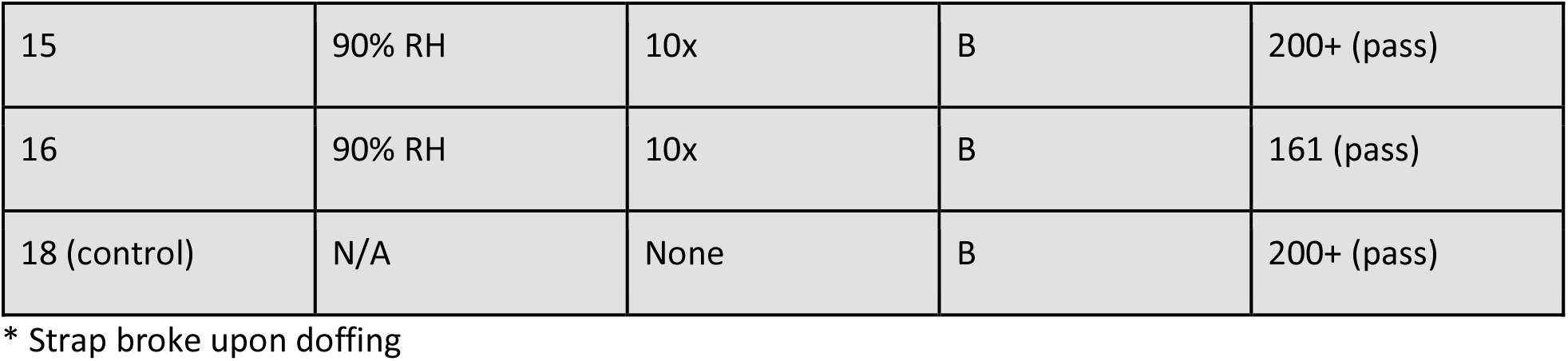
Results of quantitative fit tests

| Sample | Humidity | Heating cycles | Volunteer | Fit factor |
| --- | --- | --- | --- | --- |
| 01 | Ambient | 1x | A | 107 (pass) |
| 02 | Ambient | 1x | A | 195 (pass) |
| 03 | Ambient | 1x | A | 165 (pass) |
| 04 | Ambient | 1x | A | 149 (pass) |
| 05 | Ambient | 10x | A | 200+ (pass)* |
| 06 | Ambient | 10x | A | 200+ (pass) |
| 07 | Ambient | 10x | A | 200+ (pass) |
| 08 | Ambient | 10x | A | 200+ (pass) |
| 17 (control) | N/A | None | A | 181 (pass) |
| 09 | 90% RH | 1x | B | 200+ (pass) |
| 10 | 90% RH | 1x | B | 163 (pass) |
| 11 | 90% RH | 1x | B | 200+ (pass) |
| 12 | 90% RH | 10x | B | 200+ (pass) |
| 13 | 90% RH | 10x | B | 200+ (pass) |
| 14 | 90% RH | 10x | B | 200+ (pass) |
| 15 | 90% RH | 10x | B | 200+ (pass) |
| 16 | 90% RH | 10x | B | 161 (pass) |
| 18 (control) | N/A | None | B | 200+ (pass) |

\* Strap broke upon doffing

None of the processed samples showed any qualitative change in feel or appearance. One of the elastic straps on sample 05 snapped upon doffing the FFR after passing the quantitative fit test.

All samples subjected to dry heat cycles passed the quantitative fit tests with a fit factor of >100. After one heating cycle there was no significant change to the quantitative fit test result for the four tested samples. After 10 cycles all four tested samples had the maximum score upon fit testing. Due to FFRs 01-08 being tested sequentially, we are unable to disambiguate whether the improvement in quantitative fit test results for FFRs treated for ten cycles versus one cycle is due to the heat treatment or an improvement in donning procedure over time.

Likewise, all samples subjected to moist heat cycles passed the quantitative fit tests with a fit factor >100. There was no significant change to the quantitative fit test result for the eight samples tested, nor a correlation between score and number of treatment cycles. No significant difference was observed between the two volunteers.

In our initial experiments, we attempted to heat an FFR in an oven with an exposed heating element. We found that the FFR heated rapidly and showed visible signs of softening and melting. We believe this is due to the infrared radiation emitted by heating elements, which typically operate at temperatures of ∼800-1000 ºC. Polymers strongly absorb blackbody radiation (2-3 μm wavelength) emitted by the heating elements at this temperature. We therefore caution against using any heating method which exposes the FFR directly to radiation from the heat source.

While we found that the quantitative fit factor of FFRs was not affected by up to ten heating cycles to 75 ºC, a recent report by Fischer et. al. found that the quantitative fit factor of FFRs was only retained for 2 cycles of 70 ºC. The key difference in the two studies was the treatment of samples between heating cycles. In the present study, we donned and doffed the FFR a single time but did not simulate donning and doffing in between heating cycles. In Fischer et. al., the FFR was donned and worn for two hours in between cycles. This suggests that use duration and number of donning/doffing cycles of the FFR, perhaps even independent of heat treatment (or other decontamination protocol such as VHP), may play an important role in the quantitative fit factor of FFRs that are utilized beyond the recommended single-use. Additional studies are needed to disambiguate the effects of total use duration, donning/doffing cycles, and decontamination protocol on the quantitative fit factor of FFRs and in particular on the elastic head and neck straps.

### Viral activity results

Dry heat inactivation of MHV, a coronavirus previously used as a SARS-CoV surrogate virus for validating decontamination protocols,^12^ was used to confirm that heating to 75ºC for 30 min inactivates high titers of coronavirus on FFR filter material. Diluting MHV in media containing 10% FBS prior to inoculating the FFR filter material simulated the presence of proteins found in respiratory secretions that may increase viral resistance to inactivation. ^12^ No viral activity was detected after the heat treatment. TCID50 measurements on the coupons used in these studies show ≥ 6 log reduction after heating to 75 ºC for either 30 or 60 minutes as compared to room temperature activity for similar time periods as seen in Figure 2. These results are generally consistent with a recent report that 70 ºC dry heat for 60 minutes inactivates SARS-CoV-2 on FFR filter material (≥ 4 log reduction)^6^ and provide further evidence that dry heat provides relatively rapid inactivation of coronaviruses on FFR filter material, especially as compared to untreated FFRs stored for the same amount of time at room temperature.

**Figure 2:**
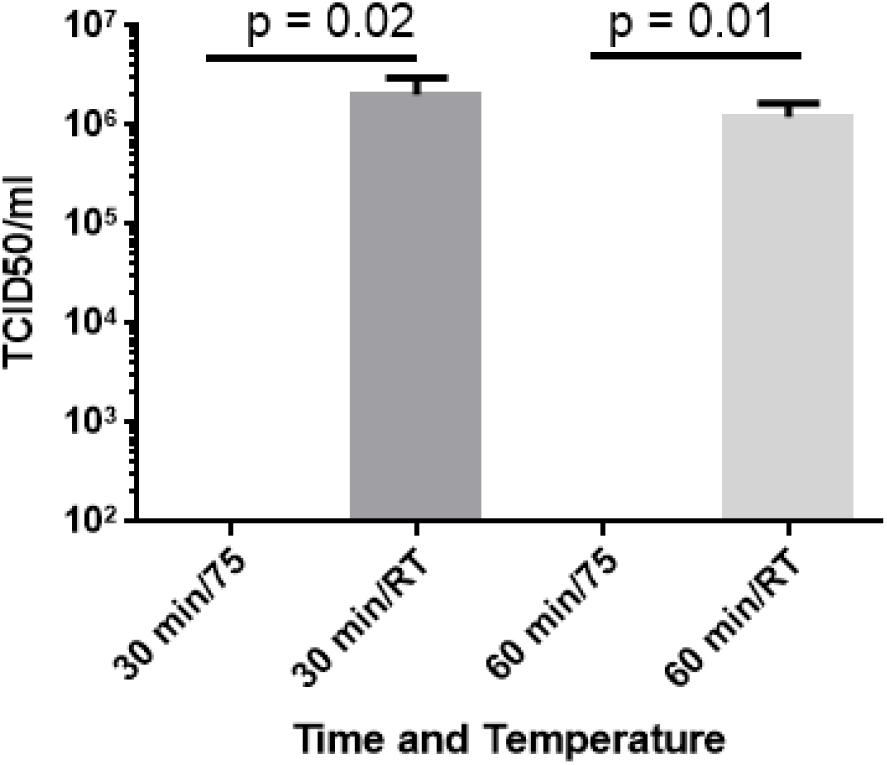
Viral activity after 30 or 60 minutes of dry heat (75 ºC) and after 30 or 60 minutes at room temperature (RT). Heated samples showed a ≥ 6 log reduction in viral activity.

## Conclusion

We subjected N95 FFRs to one and ten heating cycles up to 75 ºC under dry and humid (90% relative humidity) conditions. Quantitative fit testing did not show any degradation in the fit factor, showing that the form and fit of these FFRs was retained following the heat treatments. We also found that dry heating to 75 ºC reduced the viral activity of MHV on an FFR filter element by ≥ 6 log. These temperatures and times have already been shown to inactivate other coronaviruses (including SARS-CoV-2) in liquid media^7,8,9^ and FFR filter material,^6^ and our study provides further evidence that virus dried on FFR filter material can also be inactivated at by heating. Previous studies have shown that these temperatures have do not to negatively impact filter efficiency and airflow of melt-blown propylene filter elements found in N95 FFRs. ^3,4^

The emerging evidence shows that heat treatments may be used as an effective method for re-using N95 FFRs. It should be noted that heat treatment is not a broad-spectrum sterilant and that N95 FFRs are normally meant for one-time use. However, in emergency situations heat treatments specifically to inactivate coronaviruses may be developed using commonly available equipment (incubators, blanket warmers, clothes dryers, ovens, etc.). Heat treatments may therefore serve as a rapid method for re-use of FFRs in areas where FFRs are in critically short supply, specialized decontamination equipment (e.g., VHP) is not available, and surface sterilization (e.g., ultraviolet germicidal irradiation (UVGI)) is insufficient. However, important questions remain on the retention of fit factor after long-term use and repeated donning/doffing cycles to help resolve conflicting data on the number of cycles for which quantitative fit factor can be maintained.

## Data Availability

Data may be made available on request.

## Acknowledgment

This work was performed under the auspices of the U.S. Department of Energy by Lawrence Livermore National Laboratory under Contract DE-AC52-07NA27344. We kindly thank Dr. Susan Weiss at University of Pennsylvania Perelman School of Medicine for providing MHV strain A59.

## Footnotes

1 See summary on the CDC website, https://www.cdc.gov/coronavirus/2019-ncov/hcp/ppe-strategy/decontamination-reuse-respirators.html (retrieved 4/14/20)

2 For examples, see FDA letter to STERIS Corporation, https://www.fda.gov/media/136843/download and FDA letter to Battelle Memorial Institute, https://www.fda.gov/media/136529/download (retrieved 4/14/20)

3 Viscusi, et. al., *The Annals of Occupational Hygiene*, **53**, 815 (2009)

4 Liao, et. al., MedRxiv pre-print, https://doi.org/10.1101/2020.04.01.20050443

5 Kumar, et. al., MedRxiv pre-print, https://doi.org/10.1101/2020.04.05.20049346

6 Fischer, et. al., MedRxiv pre-print, https://doi.org/10.1101/2020.04.11.20062018

7 Rabenau, et. al., *Medical Microbiology and Immunology*, **194**, 1 (2005)

8 Darnell, et. al., *Journal of Virological Methods*, **121**, 85 (2004)

9 Chin, et. al., *The Lancet Microbe*, **in press** (2020). https://doi.org/10.1016/S2666-5247(20)30003-3

10 https://www.osha.gov/laws-regs/regulations/standardnumber/1910/1910.134AppA (retrieved 4/14/20)

11 Hierholzer and Killington, “Virus isolation and quantitation” in *Virology Methods Manual* (Mahy and Kangro, eds), 25–47 (1996)

12 Casanova, et. al., *Appl. and Environ. Microbiology*, **76**, 2712 (2010)

